# Validation of a urine- based proteomics test to predict clinically significant prostate cancer: complementing MRI pathway

**DOI:** 10.1101/2024.04.16.24305475

**Authors:** Maria Frantzi, Ana Cristina Morillo, Guillermo Lendinez, Ana Blanca-Pedregosa, Daniel Lopez Ruiz, Jose Parada, Isabel Heidegger, Zoran Culig, Emmanouil Mavrogeorgis, Antonio Lopez Beltran, Marina Mora-Ortiz, Julia Carrasco-Valiente, Harald Mischak, Rafael A Medina, Juan Pablo Campos Hernandez, Enrique Gómez Gómez

**Affiliations:** Mosaiques diagnostics GmbH, Department of Biomarker Research, Hannover, Germany; Reina Sofía University Hospital, Maimonides Institute of Biomedical Research of Cordoba (IMIBIC), University of Cordoba (UCO), Department of Urology, Cordoba, Spain; Virgen del Rocio University Hospital, IBIS, Department of Urology, Seville Spain; Reina Sofía University Hospital, Maimonides Institute of Biomedical Research of Cordoba (IMIBIC), University of Cordoba (UCO), Department of Radiology, Cordoba, Spain; Virgen del Rocio University Hospital, IBIS, Department of Radiology, Seville Spain; Medical University of Innsbruck, Department of Urology, Innsbruck, Austria; Institute for Molecular Cardiovascular Research (IMCAR), RWTH Aachen University Hospital, 52074 Aachen, Germany; University of Cordoba, Department of Pathology, Cordoba, Spain; Lipids and Atherosclerosis Unit, Internal Medicine Unit, Reina Sofía University Hospital, Maimonides Institute of Biomedical Research of Cordoba (IMIBIC), University of Cordoba (UCO), Cordoba, Spain; University of Glasgow, Institute of Cardiovascular and Medical Science, Glasgow, United Kingdom

## Abstract

**Purpose:** Prostate cancer (PCa) is the most frequently diagnosed cancer in men. One major clinical need is to accurately predict clinically significant PCa (csPCa). A proteomics based 19-biomarker model (19-BM) was previously developed using Capillary Electrophoresis-Mass Spectrometry (CE-MS) and validated in 1000 patients at risk for PCa. Here, our objective was to validate 19-BM in a multicentre prospective cohort of 101 biopsy-naive patients using current diagnostic pathways.

**Materials and Methods:** Urine samples from 101 PCa patients were analysed through CE-MS. All patients underwent MRI using a 3-T system. The 19-BM score was estimated via a support vector machine-based software (MosaCluster; v1.7.0), employing previously established cut-off criterion of -0.07. Previously developed diagnostic nomograms were calculated along with MRI.

**Results:** Independent validation of the 19-BM yielded a sensitivity of 77% and specificity of 85% (AUC:0.81). This performance surpasses that of PSA (AUC:0.56), and PSA density (AUC:0.69). For PI-RADS≤ 3 patients, the 19-BM showed a sensitivity of 86% and specificity of 88%. Integrating the 19-BM with MRI resulted in significantly better accuracy (AUC:0.90) compared to the individual investigations alone (AUC_19BM_=0.81; p=0.004 and AUC_MRI_:0.79; p=0.001). Examining the decision curve analysis, the 19-BM with MRI surpassed other approaches for the prevailing risk interval from 30% cut-off.

**Conclusions:** 19-BM exhibited favourable reproducibility for prediction of csPCa. In PI-RADS≤3 patients the 19-BM correctly classified 88% of the patients with insignificant PCa at the cost of one csPCa patient that was missed. Utilising 19-BM test could prove valuable complementing MRI and reducing the need for unnecessary biopsies.

## INTRODUCTION

Prostate cancer (PCa) stands as the most diagnosed cancer among men, with nearly 1.5 million new cases reported worldwide ^1^. Incidence rates are notably elevated in Europe and the United States ^2^, largely due to widespread prostate-specific antigen (PSA) testing combined with an aging population ^3^. Despite its prevalence, PCa boasts a significant survival rate, with 78% of individuals surviving ten or more years post-diagnosis^3^. This is because a considerable portion of cases represent low-risk, indolent disease unlikely to progress to lethal stages.

The diagnosis of PCa relies on histopathological confirmation via prostate biopsy cores, which follows positive results from i) digital rectal examination (DRE), ii) elevated PSA levels, and, more recently, iii) suspicious findings on multiparametric or biparametric magnetic resonance imaging (MRI) ^3^. However, PSA lacks specificity, with only about 40% of patients with elevated PSA serum levels (≥4 ng/ml) confirmed with PCa on biopsy ^3^. DRE, on the other hand, is subjective and may yield transient findings^3^. Additionally, MRI’s specificity is compromised ^4^, leading to a high prevalence of false-positive lesions needing a biopsy, compounded by inter-reader variation ^5^. At the same time, the widespread adoption of MRI in diagnosis poses challenges for radiology and urology departments due to limited MRI capacity and/or insufficiently trained personnel. Addressing this bottleneck requires a more effective entry test than PSA before an MRI. Thus, presently, the management of PCa faces a critical challenge: the lack of precise biomarker tests and diagnostic procedures to differentiate between clinically significant (csPCa) and insignificant PCa cases (insPCa) during the early stages of the disease. This distinction is crucial as only csPCa warrants diagnosis and subsequent treatment. To mitigate overdiagnosis and overtreatment, particularly for slow-growing PCa, there is a pressing need for better guidance of invasive biopsies through non-invasive means ^6^.

Single- or multi-biomarker assays based on urinary analysis, such as prostate cancer antigen-3 (PCA3) ^7^, SelectMDx ^8^, Mi-Prostate Score ^9^, and ExoDx, ^10^ have emerged but are yet to be incorporated into clinical guidelines and/ or healthcare systems ^3^. These tests typically involve sampling procedures following DRE, assuming that prostatic secretions contain valid biomarkers. To enhance patient comfort and convenience, our study suggests transitioning to biomarker investigations in urine samples collected without prior intervention.

Previously, a first proteomics study including more than 800 patients, focused on biomarkers discriminating csPCa via Capillary Electrophoresis coupled to Mass Spectrometry (CE-MS) was performed ^11^. CE-MS offers high sensitivity and resolution in profiling the proteomic content of urine ^12^. Based on previously published data, 19 peptide biomarkers were integrated into a machine learning model that was developed to predict csPCa. In an independent validation study, including 147 PCa patients, the 19-biomarker model (19-BM) was further validated resulting in an accuracy of 81% ^13^. Our aim in this study was to validate the predictive capacity of the 19-BM in a multicentre cohort comprising 101 biopsy-naive patients, utilizing contemporary diagnostic pathways, including a side-by-side comparison with MRI.

## PATIENTS AND METHODS

### Participants and study characteristics

This study adhered to the REMARK Reporting Recommendations ^14^ and followed the guidelines for biomarker identification and reporting in clinical proteomics ^15^. This validation study focused on patients suspected of having prostate cancer (PCa), who underwent an MRI and a transrectal ultrasound guided prostate biopsy at the Urology departments of Reina Sofia Hospital, Cordoba, and Virgen del Rocio University Hospital, IBIS, in Seville Spain, between June 2021 and July 2022. Ethical approval was obtained from the Reina Sofia Hospital Research Ethics Committee (**approval number PI22/01769)**, and informed consent was obtained from all participants. The patient cohort comprised individuals referred to the urology clinic of Reina Sofia Hospital or Virgen del Rocio University Hospital (IBIS) for a prostate biopsy based on clinical indications such as suspicious findings on digital rectal examination (DRE), PSA levels >10 ng/mL, or PSA levels between 3-10 ng/mL (if the free PSA ratio is low). Prospective collection of urine before biopsy was rutinely performed for biobank and a retrospective selection of those patients with no previous biopsy and MRI of sufficient quality was carried out (**Figure 1**). For transrectal prostate biopsy, systematic 12 core biopsy was performed in those with no suspicious lesion in MRI and target plus systematic biopsy in those with suspicious lesion. A total of 101 patients were included in the study, each with complete records for all primary variables, including MRI results, PSA levels, prostate volume, DRE findings, and pathology results. Biopsy specimens were examined by a urologic pathologist following the International Society of Urological Pathology 2019 modified criteria ^16^. Summary characteristics of the patient cohort are presented in **Table 1**.

**Figure 1:**
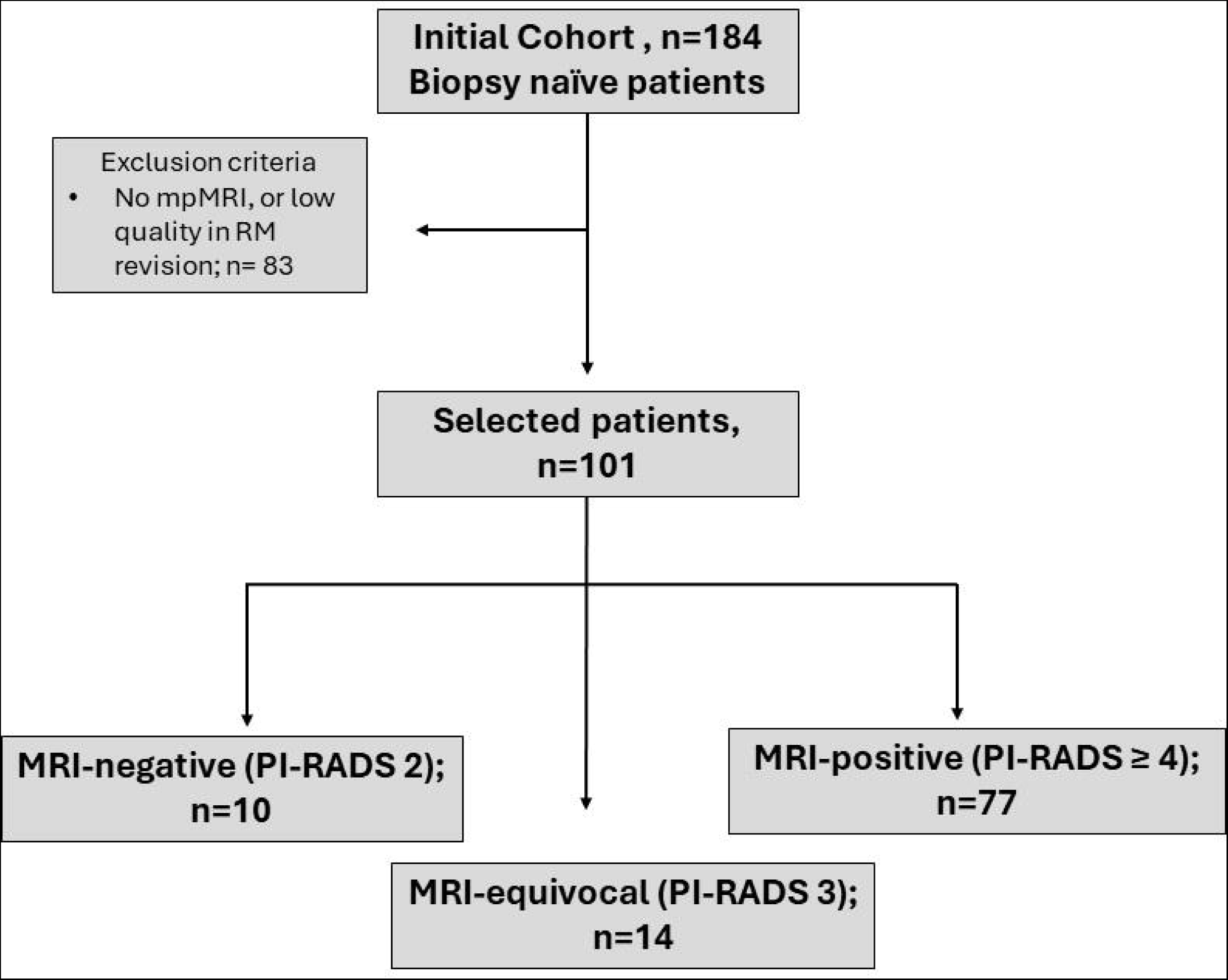
Flow diagram that visually summarises the screening process for this study focusing on biopsy-naïve men, suspicious of the presence of csPCa.

**Table 1.** Clinical and biochemical variables for the 101 patients, according to guidelines for reporting of figures and tables for clinical research in urology ^26^.

| Variable | Value |
| --- | --- |
| Number of peptides (mean (SD)) | 4202.5 (1193.4) |
| Age (mean (SD)) | 64.9 (7.9) |
| PSA (ng/ml); (median [IQR]) | 6.0 [4.5, 7.5] |
| Prostate volume (cc); (mean (SD)) | 54.3 (28.6) |
| PSAD; (median [IQR]) | 0.1 [0.1, 0.2] |
| ERSPC sig (v3; DRE); (median [IQR]) | 5.0 [3.0, 13.0] |
| DRE (%) |  |
| Normal | 78 (77.2) |
| Pathological | 18 (17.8) |
| Not performed | 5 (5) |
| PI-RADS (%) |  |
| 1 | 1 (1.0) |
| 2 | 9 (8.9) |
| 3 | 14 (13.9) |
| 4 | 43 (42.6) |
| 5 | 34 (33.7) |
| Detected PCa Tumours (%) | 73 (72.3) |
| Significant PCa tumours (ISUP ≥ 2) | 60 (59.4) |
| ISUP (%) |  |
| ISUP1 | 13 (17.8) |
| ISUP2 | 25 (34.25) |
| ISUP3 | 25 (34.25) |
| ISUP4 | 6 (8.2) |
| ISUP5 | 4 (5.5) |
| **Total number of patients** | **101** |

### Prostate MRI scan

Prostate MRI examinations were performed on a 3T MRI system (MagnetomVida, Siemens AG, Erlangen, Germany) using a 18-channel phased-array body coil and 3T MRI system (Ingenia, Philips, Eindhoven, Netherlands) using a 32-channel phased-array body coil. The protocol included T1-weighted images (T1WI), T2-weighted images (T2WI) in Three orthogonal planes, diffusion-weighted images (DWI), and dynamic contrast-enhanced images (DCE) (the last sequence was only performed at Reina Sofia Hospital). Findings and lesions were described and localized according to prostate sectors recommended by PI-RADS v 2.1 and a final assessment category was described for each patient. Images were evaluated using a PACS (Vue PACS, Philips) and Syngo.via, MRI prostate, Siemens.

### Urine collection, storage and proteomics analysis

All urine samples were meticulously collected before the prostate biopsy, aligning with established clinical protocols, and without preceding digital rectal examination (DRE). Voided urine specimens were carefully obtained using sterile containers and promptly preserved at -20°C until subsequent processing. Sample preparation was carried out as described in the discovery and prior validation studies. Details are provided in the **Supplementary Text.** CE-MS analysis and data processing was performed according to ISO13485 standards yielding quality controlled urinary datasets ^17^. A detailed description is given at the **Supplementary Text.**

### Model scoring and statistical analysis

To characterize the patient cohort, we calculated proportions, means, standard deviations, medians, and interquartile ranges for the different variables, providing a comprehensive summary in **Table 1**. The scores generated by the biomarker model were computed using support vector machine (SVM)-based software, specifically MosaCluster (version 1.7.0), as outlined in previous studies ^11^. Sensitivity and specificity for the SVM-based peptide marker pattern were calculated based on the number of correctly classified samples, as defined by biopsy, considering the previously reported cut-off criterion of (-0.07). Receiver operating characteristic (ROC) plots and the respective confidence intervals (95%CI) were based on exact binomial calculations and were calculated in MedCalc 12.7.5.0 (Mariarke, Belgium). The area under the ROC curve (AUC) was evaluated to estimate the overall accuracy independent upon a particular threshold^18^, and the values were then compared using DeLong tests. Statistical comparisons of the classification scores between the PCa risk groups and GS groups were performed by the Kruskal-Wallis rank sum test using MedCalc 12.7.5.0 (Mariakerke, Belgium)^19^. The diagnostic nomograms of 19-BM in combination with clinical variables and/or MRI were established using multiple linear regression analyses. Decision curve analysis (DCA) ^20^ examined the potential net benefit of using the diagnostic nomograms in the clinic, according to which a net benefit is defined as a function of the decision threshold at which one would consider obtaining a biopsy.

## RESULTS

### Validation of 19-BM in biopsy naive patients

We initiated the validation process by assessing the performance of the previously published 19-BM [13], following the recommendations for biomarker identification and reporting in clinical proteomics [15]. In a cohort consisting of 101 patients, 41 patients were confirmed with no PCa presence (benign conditions) or with insPCa, while 60 patients were diagnosed with csPCa (**Figure 1**; **Table 1**). Out of 101 patients, 10 were receiving a PI-RADS 2 result, 14 a PI-RADS 3 and 77 a PI-RADS ≥ 4. The proportion of any PCa type (**Figure 2a**) within PI-RADS 2, 3 and ≥ 4 was 66.7%, 55.6% and 83.1%, while the incidence of csPCa within PI-RADS 2, 3 and ≥ 4 was 25%, 40% and 70.1% respectively (**Figure 2b**). Distribution plots of the patients with benign conditions and any type of PCa (ISUP ≥ 1) according to prostate volume **(Supplementary Figure SF1a**), PSA density (PSAD; **Supplementary Figure SF1b**) and ERSPC-3 **(Supplementary Figure SF1c**) are provided across the PI-RADS groups. The estimated AUC for the 19-BM (AUC_19-BM_) to be 0.81, with a 95% confidence interval (CI) ranging from 0.72 to 0.88 (p < 0.0001; **Figure 3a**). At the validated cut-off level of -0.07, the sensitivity was calculated to be 76.7%, with a specificity of 85.4%. The 19-BM accurately classified 24 out of 28 patients that were eventually confirmed with benign lesions, 10 out of 13 patients with insPCa (ISUP 1) and 47 out of 60 patients with csPCa (ISUP ≥ 2). Considering a prevalence rate of 30% for csPCa ^21-23^, the Negative Predictive Value (NPV) for detecting csPCa was computed at 89.6%, while the Positive Predictive Value (PPV) stood at 68.8%. Furthermore, the 19-BM exhibited significant discrimination between csPCa and insPCa (p< 0.0001, Kruskal-Wallis test, **Figure 3b**), across PI-RADS (**Figure 4a**) and the ISUP groups (**Figure 4b**).

**Figure 2:**
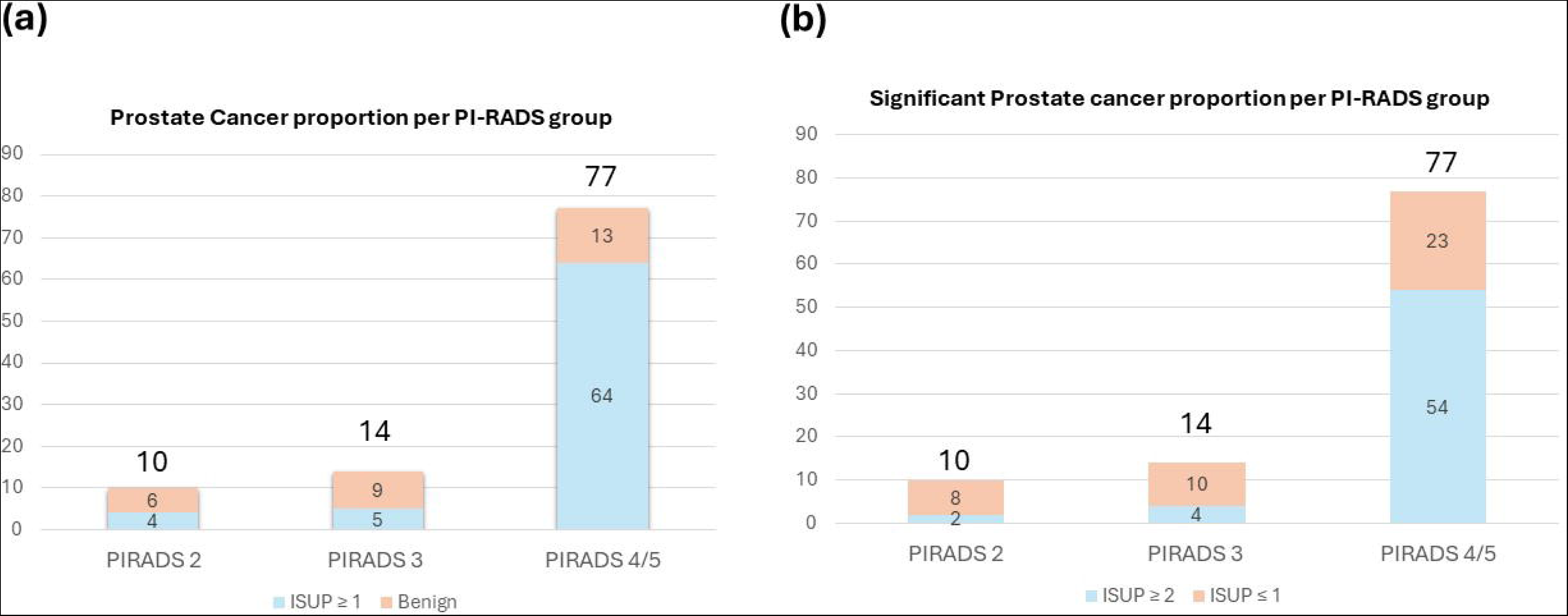
**a)** Proportions of any type of PCa (ISUP ≥ 1) and, **b)** csPCa (ISUP ≥ 2) across PI-RADS groups; distribution plots of **c)** prostate volume, **d)** PSAD, and **e**) ERSPC estimates across PI-RADS groups. In blue were the patients that were confirmed with benign prostate pathologies and in red were those confirmed with PCa (ISUP ≥ 1).

**Figure 3:**
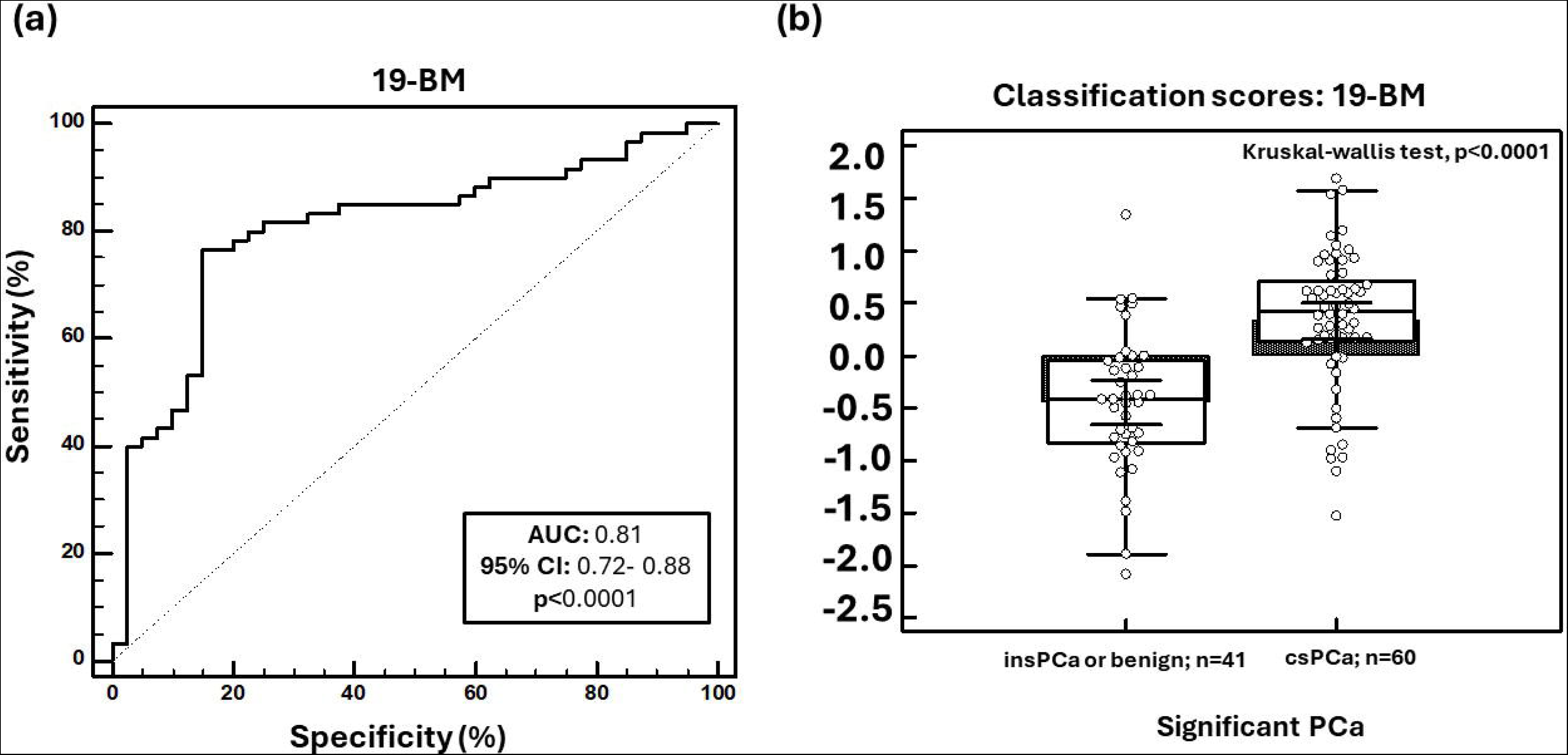
**a)** Receiver operating characteristics (ROC) analysis displaying the performance of the 19-BM for predicting csPCa; **b)** Classification scores, presented in Box-and-Whisker plots grouped according to the csPCa (n=60) and insPCa (n=41). A post-hoc rank test was performed using Kruskal-Walli’s test.

**Figure 4:**
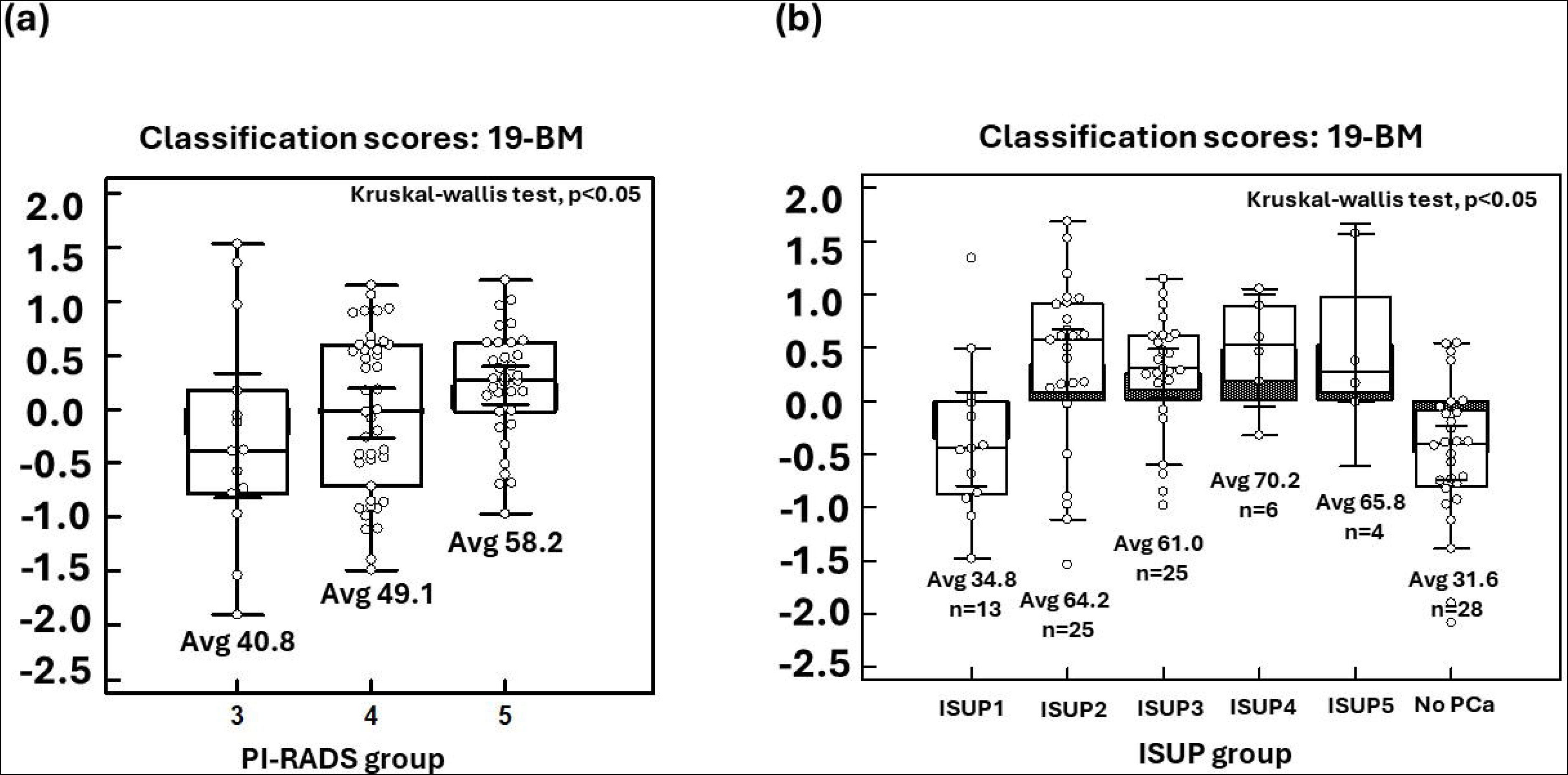
**a)** Classification scores displaying the level of discrimination across the different PI-RADS groups and **b)** across ISUP grading groups. A post hoc rank test was performed using the Kruskal-Wallis test.

### Comparative analysis with PSA and PSAD

Subsequently, we conducted a comparative analysis between serum PSA levels, PSAD and the 19-BM to discern their efficacy in predicting csPCa. Multivariate analysis revealed that the 19-BM surpassed PSA, with an AUC for PSA of 0.56 (95% CI: 0.46 - 0.66; p = 0.0019; **Figure 5a** and **5b**). Remarkably, the 19-BM detected 6 out of 8 PCa patients who were missed by serum PSA, all of them bearing a csPCa. Furthermore, both the NPV and PPV estimates based on the 19-BM were superior to those based on PSA, with NPV at 79.6% and PPV at 30.4%. Similarly, when considering PSAD, the 19-BM outperformed PSAD in predicting csPCa, with an AUC of 0.69 (95% CI: 0.58-0.77; p = 0.04; **Figure 5a** and **5b**). The sensitivity of PSAD was estimated at 73.3% (95% CI: 60.3 - 83.9%), while the specificity was 63.4% (95% CI: 46.9 – 77.9%; PSAD ≥ 0.1). Ten patients with csPCa were detected by the 19-BM but were missed when considering PSAD. PPV and NPV estimates for PSAD were once more inferior compared to 19-BM, resulting in 45.8% and 84.5%, respectively.

**Figure 5:**
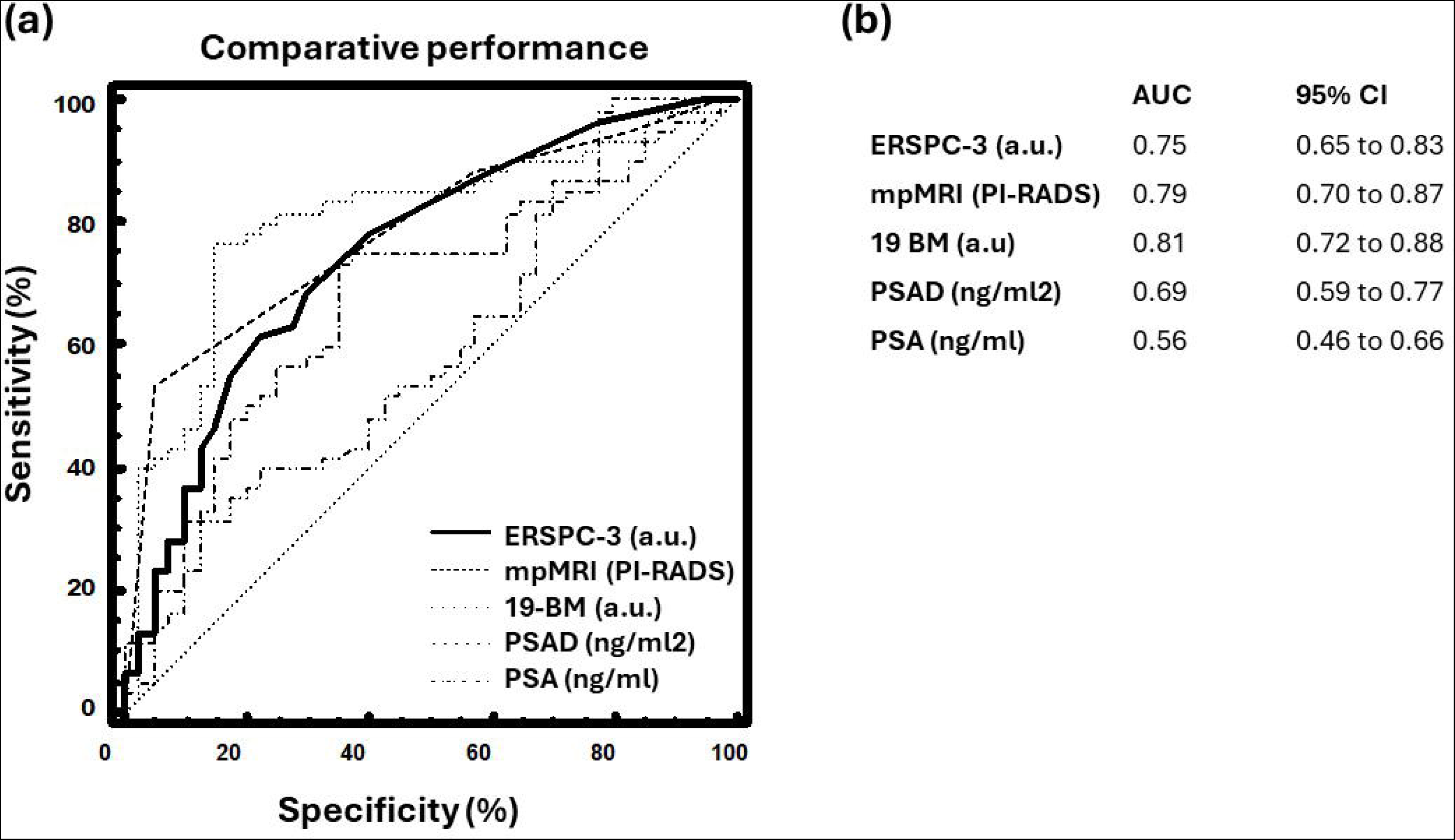
**a)** Comparative analysis depicted by receiver operating characteristics (ROC) curves of the 19-BM with serum PSA measurements, PSAD, ERSPC-3, and MRI; **b**) Information about the AUC estimates and 95% confidence intervals.

### Comparison of 19-BM with the ERSPC clinical risk calculator

To evaluate whether the 19-BM could enhance current state-of-the-art clinical prognosticators, we further compared the 19-BM performance to the estimates derived from the European Randomized Study of Screening for Prostate Cancer (ERSPC) risk calculator for detecting high-risk prostate cancer (ERSPC-3 with DRE), as depicted in **Figure 5a**. The performance of the 19-BM exceeded that of the ERSPC-3 risk calculator (AUC_ERSPC-3_ = 0.75; 95% CI: 0.65-0.83). At the cohort optimal cut-off level (ERSPC-3 ≥ 6), the sensitivity of ERSPC-3 was 61.7% (95% CI: 48.2-73.9%) with a specificity of 78.1% (95% CI: 62.4 - 89.4%). ERSPC-3 missed 22 patients with confirmed csPCa, 17 of whom were correctly identified by the 19-BM. NPV and PPV for ERSPC-3 were estimated at 54.7% and 82.7% respectively, both lower than the ones from 19-BM. Similarly, a direct comparison was performed between the 19- BM and the MRI. MRI showed similar AUC as 19-BM (AUC_MRI_: 0.79; 0.71-0.87; 95% CI; **Figures 5a** and **5b**). Considering PI-RADS ≥ 4 (as positive), the sensitivity of MRI was 90% (79.5 to 96.2%), while the specificity only 43.9 (28.5 to 60.3%). NPV and PPV estimates were estimated at 91.1% and 40.8%, respectively.

### Validation of published nomogram and value within equivocal MRI

Like our previous study [13], we integrated PSAD and age with the 19-BM to validate the previously developed diagnostic nomogram (19-BM, age and PSAD). This integration resulted in a slightly improved but non-significant AUC value of 0.82 (95% CI: 0.73-0.89; **Figure 6a**). At the previously defined optimal cut-off for the diagnostic nomogram (>0.1766), the sensitivity in this study is estimated at 96.7% (95% CI: 88.5-99.6%) and the specificity at 26.8% (95% CI: 14.2-42.9%). The NPV for detecting csPCa has been computed at 95.5%, while the PPV at 36.3%. Within PI-RADS ≤3 patient group the 19-BM resulted in an AUC of 0.82 (0.62 to 0.95; 95% CI; **Figure 6b**), with 85.7% sensitivity and 88.2% specificity, similar to the accuracy of the integrative nomogram (19-BM, age and PSAD; AUC: 0.80; 0.59 to 0.93; 95% CI), that resulted in estimated sensitivity of 86% and specificity of 76.5%.

**Figure 6:**
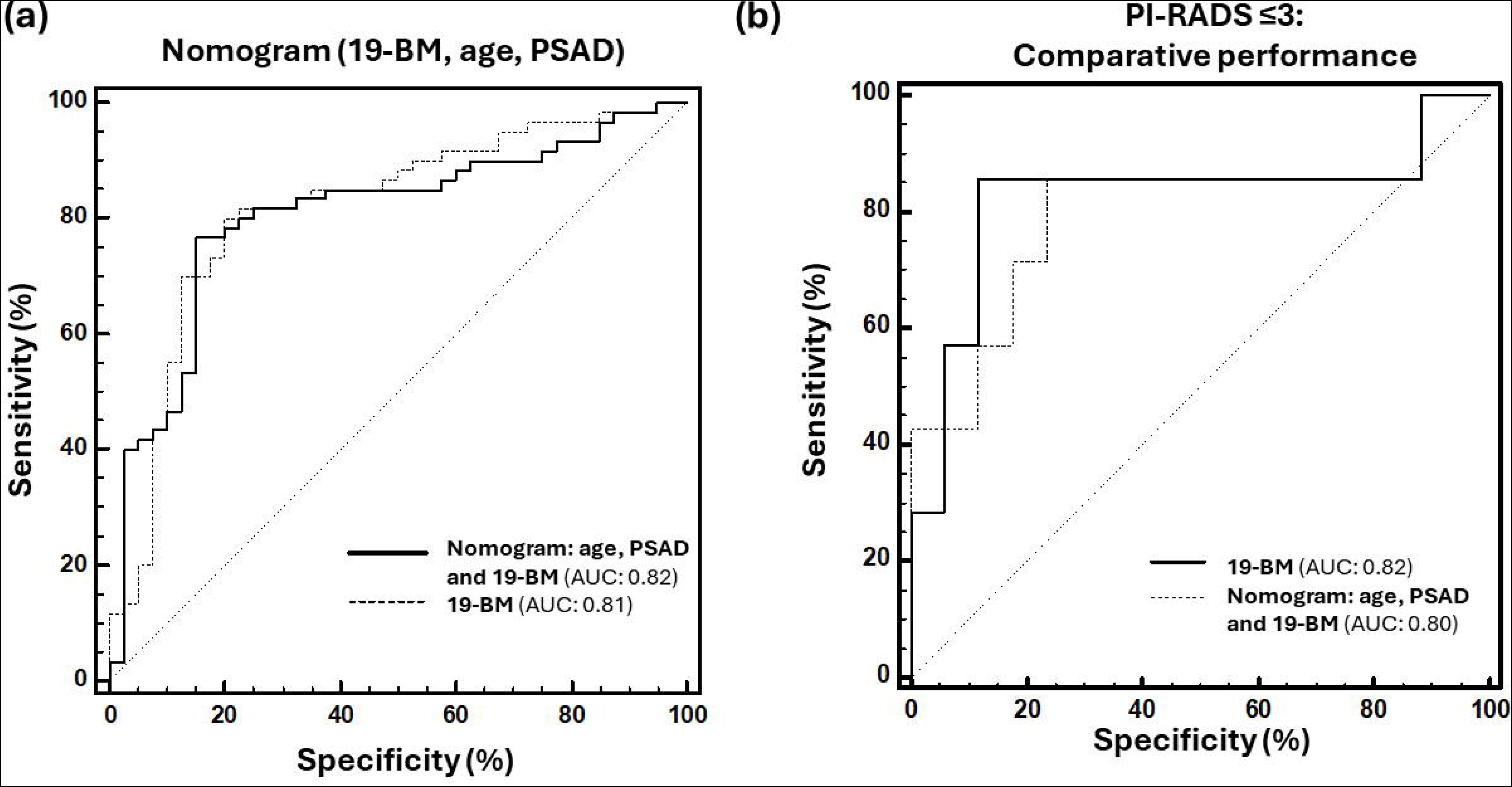
**a)** ROC analysis displaying the performance of the previously published nomogram integrating age, 19-BM and PSAD; **b**) Accuracy of both 19-BM and the nomogram is provided within the equivocal MRI results.

### Added value: integration of 19-BM with MRI

The added value of the 19-BM test was also investigated in addition to MRI, to investigate benefit upon integration of both tests. A significantly superior performance reached 90% (AUC: 0.90; 0.83 to 0.95; 95% CI; **Figure 7a**), resulting in a sensitivity estimation of 88% and specificity of 78%. NPV and PPV values were estimated at 93.8% and 63.2% respectively. To evaluate the clinical benefit of the integrative model (19-BM and MRI), we performed a decision curve analysis also in comparison with the rest single tests. The results indicated a clear benefit of the integrative model, particularly in the middle range of the risk thresholds (20-60%; **Figure 7b**).

**Figure 7:**
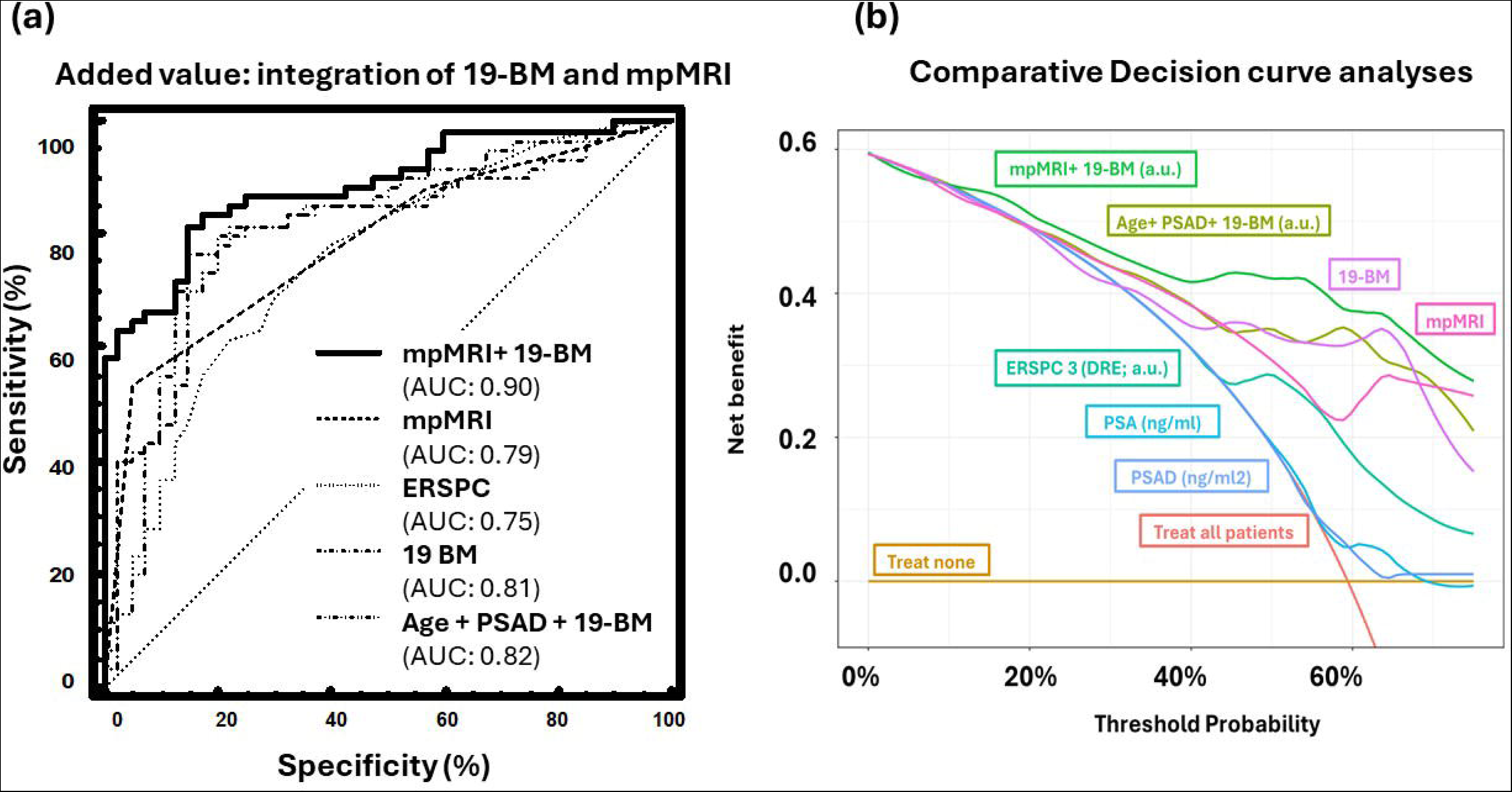
**a)** Added value for 19-BM upon integration with MRI; **b)** Results of the decision curve analysis, comparing the net benefit for the prediction of csPCa on biopsy.

## DISCUSSION

Despite improving the diagnosis pathway, there is still a high number of clinically insPCa that is detected through a substantial number of biopsies mainly driven by false positive or equivocal MRI lesions. Incorporating a non-invasive biomarker test like 19-BM with enhanced accuracy into a diagnostic workflow alongside MRI, has the potential to improve on over-diagnosis, particularly to potentially delay or even avoid regular biopsies for accurately detecting csPCa. Studies based on high resolution proteomics (like CE-MS) have been published predicting PCa biopsy outcome with aim to guide prostate biopsy. Through the use of CE-MS proteomics, a biomarker model based on 19 urinary peptides (19-BM) was previously established with the aim to accurately detect csPCa in 823 patients suspicious for PCa (reporting an AUC of 0.81, outperforming PSA and ERPSC) ^11^. Building upon the previous report and considering urine collection without any prior prostate massage or DRE, in this study, we aimed to validate the 19-BM in biopsy-naïve patients at high risk for csPCa. This involved conducting a side-by-side comparison with MRI.

Previous studies demonstrated an accuracy of 81% which is fully reproduced in this study. In this study, 19-BM again performed better than PSA, PSAD and ERSPC-3 and successfully classified most csPCa that were missed by them. Moreover, both the NPV of 19-BM and the PPV were also higher than these variables estimate. The previously published nomogram considering PSAD, age and 19-BM was additionally tested. The accuracy of the nomogram was once more only slightly better than the 19-BM alone. Comparing the use of PSA at 3 ng/ml for MRI referral, adopting a companion diagnostic tool, is projected to reduced benign biopsies by 64.3% and 61.5% of those that would result in a detection of insPCa (**Figure 8**). At the same time, 19-BM can accurately predict 83% of the csPCa within those patients with equivocal MRI results. In a clinical context where individuals present with high risk of prostate cancer despite low PSA levels (< 10ng/ml), such as in this study, it is relevant to emphasise on the necessity of high NPV value. The required high NPV for accurate detection of csPCa (90%) was achieved in this study. A main advantage of the 19-BM test is that unlike almost all the urinary biomarker tests, performing a DRE is not necessary for the analysis.

**Figure 8:**
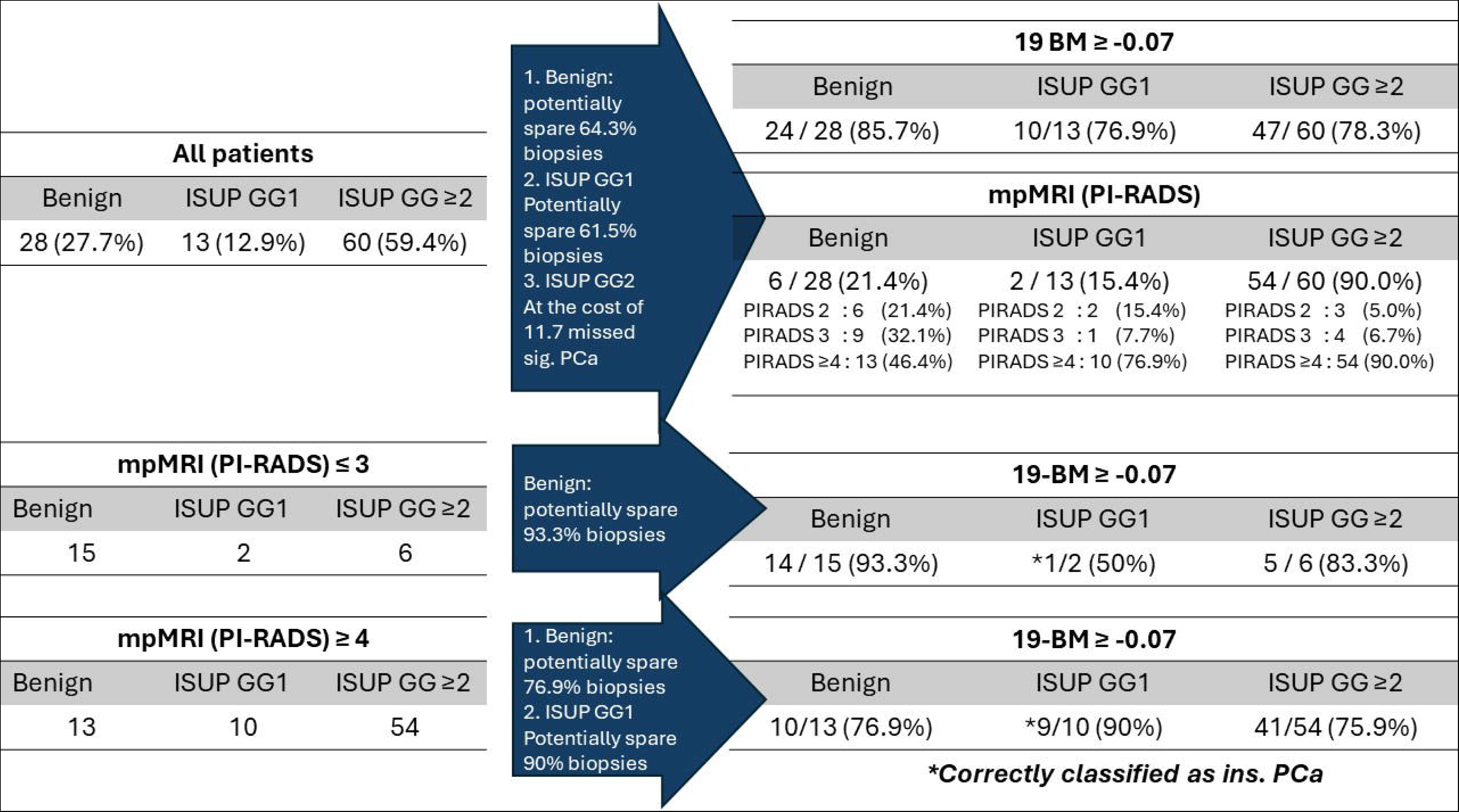
Potential reduction in the number of biopsies in different diagnostic pathway scenarios.

Direct comparison between the 19-BM and other reported markers is unfortunately not possible because of different cohorts, including different PCa detection rates. However, this study clearly shows that the 19BM is robust and consistent as it maintained its accuracy in clinical cohorts with different prevalence and diagnosis approaches. An indirect comparison with other urine biomarkers show at least the same or best performance considering and interval AUC for some of them such as Select MdX^21^ or PCA3 (0.76 and 0.73, respectively), with the advantage of urine collecting with no manipulation ^24^.

A second limitations it that MRI accuracy and detection rates could be biased as patient selection was performed in the clinic based on other variables such as PSAD, DRE, and family history to include men at high risk for PCa. Nevertheless, based on the data presented, implementation in an investigative setting seems to be highly justified. Moreover, the study includes a high risk population where the proportion of csPCa is calculated at 59.4%, higher compared to other trials like PRECISION biopsy trial ^25^. Collectively, the data presented in this study could demonstrate the utility of a multimodal approach for improved non-invasive detection of csPCa. Effective discrimination between cs and insPCa is expected to improve on reducing the number of diagnostic biopsies, and thus have a positive impact on PCa patient management, by improving patient compliance and reducing over-treatment and the associated costs. Considering, the high NPV the clinical utility of the presented nomogram could also be potentially investigated in the context of guiding MRI.

## ADDITIONAL INFORMATION

### Ethics approval and consent to participate

Ethical approval was obtained by the Ethics committee of the University Hospital of Reina Sofia (Cordoba, Spain, approval number PI22/01769; 31/08/2022), following the Declaration of Helsinki and informed consent was obtained from all participants.

## Data Availability

All data produced in the present study are available upon reasonable request to the authors

## Availability of Data and Materials

Mass spectrometry raw data can be provided upon request.

### Funding

This work was funded by PI22/01769 [FIS (Science and Innovation Ministry, ISC III, FEDER)] and ProSTRAT AI (01DS23014; Federal Ministry of Education and Research/ BMBF/ EUREKA network). Marina Mora-Ortiz has been awarded funding from the European Union’s Horizon 2020 research and innovation programme under the Marie Skłodowska-Curie grant agreement No 847468.

### Supplementary Files-Legends

**Supplementary Text:** Urine sampling and proteomics analysis.

**Supplementary Figure SF1:** Distribution plots of the patients with benign conditions and any type of PCa (ISUP ≥ 1) according to: **a)** prostate volume, **b)** PSAD, and **c**) ERSPC estimates. In blue were the patients that were confirmed with benign prostate pathologies and in red were those confirmed with PCa (ISUP ≥ 1).

