## Supplementary figures and images for "Validation of a urine- based proteomics test to predict clinically significant prostate cancer: complementing MRI pathway"

### Supplementary Figure SF1a

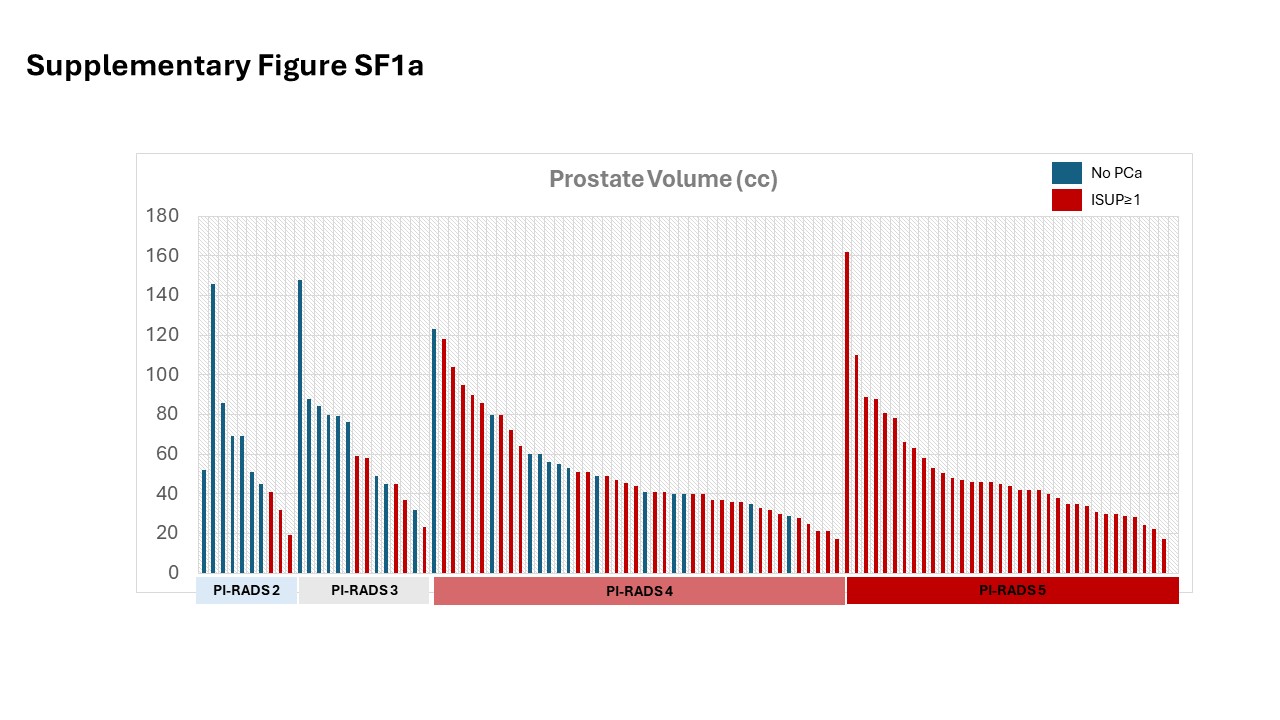

### Supplementary Figure SF1b

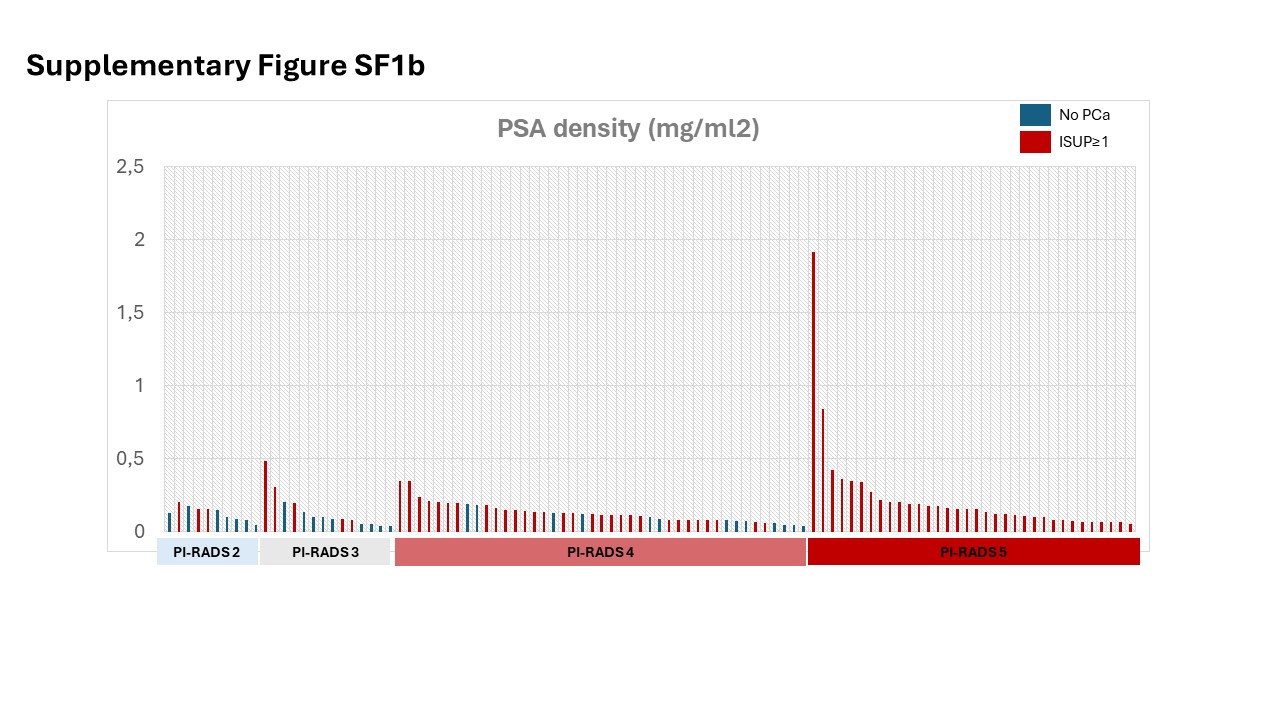

### Supplementary Figure SF1c

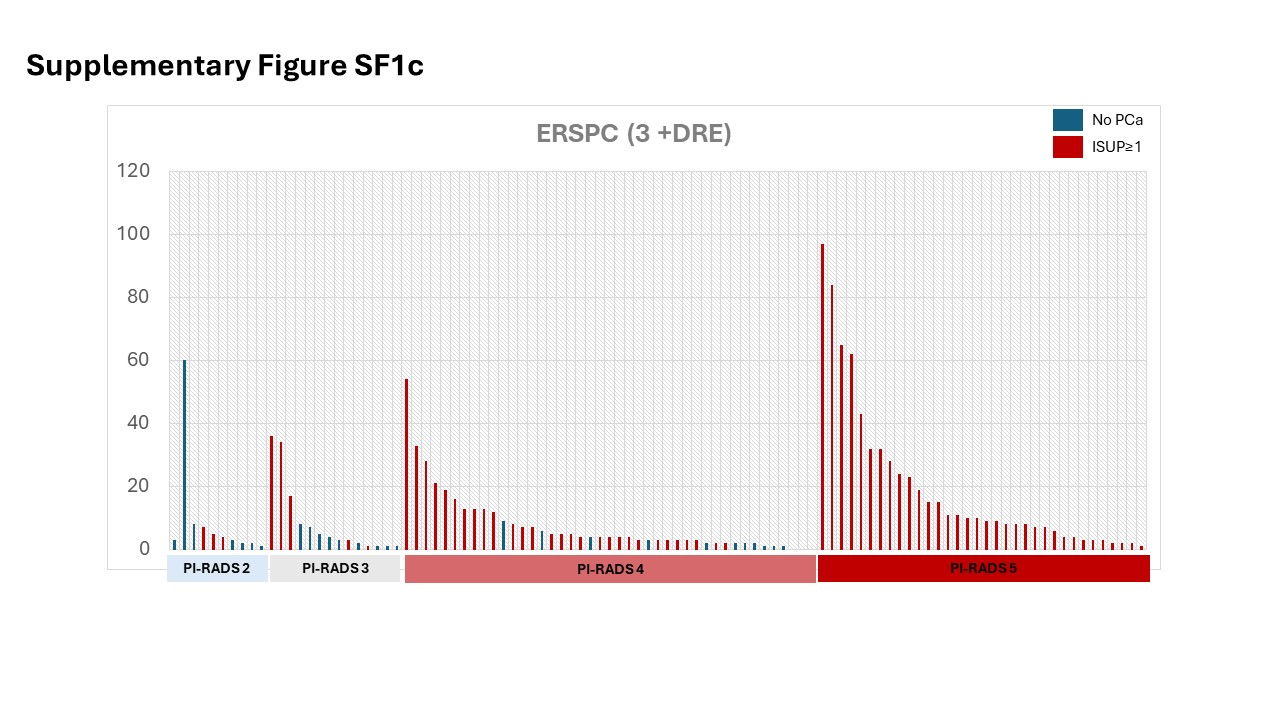
